# Trends and determinants of Acute Respiratory Infection symptoms among Under-five children in Cambodia: Analysis of 2000 to 2014 Cambodia Demographic and Health Surveys

**DOI:** 10.1101/2022.11.17.22282444

**Authors:** Samnang Um, Punleak Pin, Daraden Vang, Darapheak Chau

## Abstract

**INTRODUCTION:** Acute Respiratory Infections (ARI) is public health concern. According to the World Health Organization (WHO), ARI is responsible for 3.5% of all diseases in the world and more than 808,000 (15%) of all under-five deaths worldwide in 2017. ARI continues to be one of the leading causes of childhood morbidity and mortality in Cambodia, particularly among children under five of age. We aimed to assess the trends over time of ARI symptoms and examine the socio-demographic, behavioral, and environmental factors associated with ARI symptoms among Cambodian children aged 0-59 months across 2000, 2005, 2010, and 2014.

**METHODS:** We used existing children’s data from the Cambodia Demographic and Health Survey (CDHS) with a total of children ages 0-59 months included 7,828 in 2000, 7,621 in 2005, 7,727 in 2010, and 6,864 in 2014, respectively. All statistical estimations were carried out using STATA V16, within the survey-specific command “svy” using the standard sampling weight (v005/1,000,000), clustering, and stratification. We used simple and multiple logistic regression to determine the main predictors of ARI symptoms.

**RESULTS:** ARI symptoms in the previous two weeks in children aged 0-59 months in Cambodia decreased from 19.9% in 2000 to 8.6% in 2005 to 6.4% in 2010, then fewer at 5.5% in 2014. The main factors that increased the likelihood of ARI symptoms were children ages 6-11 months [AOR=1.91; 95% CI: 1.53-2.38], 12-23 months [AOR=1.79; 95% CI: 1.46-2.20], and 24-35 months [AOR=1.41; 95% CI: 1.13-1.76], mothers who smoke cigarette [AOR=1.61; 95% CI: 1.27-2.05], and children born into households had non-improved toilets [AOR=1.20; 95% CI: 0.99-1.46]. However, the following factors were found to be associated with decreased odds of having ARI symptoms: Mothers with higher education [AOR=0.45; 95% CI: 0.21-0.94], breastfeeding children [AOR=0.87; 95% CI: 0.77-0.98], and children born into richest wealth quantile [AOR=0.73; 95% CI: 0.56-0.95], respectively. Survey years in 2005 [AOR=0.36; 95% CI: 0.31-0.42], 2010 [AOR=0.27; 95% CI: 0.22-0.33], 2014 [AOR=0.24; 95% CI: 0.19-0.30].

**CONCLUSION:** The trends of ARI symptoms among children under five in Cambodia significantly decreased from 2000-2014. Mothers who smoke cigarettes, young children ages (6-35 months), and household unimproved toilet facilities are factors that independently increased the likelihood that children would develop ARI symptoms. Inversely, factors were found to be associated with decreased odds of having ARI symptoms: Mothers with higher education, breastfeeding children, and children born into the richest wealth quantile and Survey years. Therefore, government and child family programs must promote maternal education, particularly infant breastfeeding. The government ought to support maternal education and infant breastfeeding in the interest of early childhood care.

## INTRODUCTION

Acute respiratory infections (ARI) is public health concern, and the leading cause mortality and morbidity of children [1]. It is estimated that 5 million children under the age of five (U5) died worldwide in 2020 [2]. All of these deaths could have been avoided and treated if there had been access to more affordable health, sanitation, and hygienic interventions [2]. In Sub-Saharan Africa and Southern Asia, where there were 5 million U5, more than 80% of them were reported in 2020 [2, 3]. ARI refers to an infection of the airways, which includes the upper and lower respiratory tracts, brought on by a variety of pathogens with an evolution time of less the with an evolution of fewer than 15 days, including bacteria, viruses, fungi, and parasites. Except for those who had only a blocked nose, most children exhibit one or more ARI signs and symptoms such as coughing, difficulty breathing or dyspnea, or tachypnea [3-6]. With more than 808,000 (15%) of all deaths of U5 worldwide in 2017 attributable to ARI symptoms, this condition continues to be the leading cause of death [7]. World Health Organization (WHO), estimated 3.5% of the global disease burden is caused by ARI, and is responsible for between 30% to 50% of all pediatric outpatient visits and more than 30% of pediatric admissions in low and middle-income countries [2]. In Cambodia, ARI continue to be one of the leading causes of U5 morbidity and mortality; percentage of U5 exhibiting ARI symptoms peaked at 20% in 2000, and has since decreased to 6% in 2014 [8-10]. Due to ARI symptoms, children between 37% in 2000 and 69% in 2014 were brought to a medical facility in Cambodia [8-10]. Nearly 83% of children with symptoms of ARIs received antibiotics [8-10]. Common diseases that can be prevented and treated in U5 populations that causes of death are from diseases included ARI, Diarrheal, Malaria, and Malnutrition, respectively [8-10]. Previous research conducted in Zambia using Demographic and Health Survey (DHS) from 1996-2014 showed that children whose mothers had a secondary or higher education were less likely to have ARI symptoms (AOR=0.30) than those with no education. Children who were underweight had a higher likelihood of having ARI symptoms (AOR=1.50) than children who were not. Compared to households that used electricity, children who lived in homes where biomass fuels like charcoal and wood were used had a higher risk of ARI symptoms (AOR=2.67) [11]. Children under the age of one were more likely than older children to have ARI symptoms [11-13]. Urban areas had a higher prevalence, according to study from India [14]. Children from low-income families who lack access to healthcare services are more likely to develop ARI symptoms [4, 12-14]. Despite, the burden of ARI symptoms on morbidity and mortality in children U5 in the world, there is limited data to evaluate the trends and the risks of socio-demographic, behavioral, and environmental factors that are mainly associated with ARI symptoms among children aged 0-59 months in Cambodia. The availability of data on trends and risk factors of ARI symptoms is vital because achieving Sustainable Development Goal on improving health and wellbeing will depend on the existing efforts to prevent and control ARIs in all WHO regions [15]. Thus, this study aimed to describe the trends over time of ARI symptoms in children aged 0-59 months across 2000, 2005, 2010, and 2014 CHDS and looked at the relationships between socio-demographic, behavioral, and environmental factors with ARI symptoms in children under five of age in Cambodia.

## METHODS

### DATA SOURCE

We used existing children’s data from the 2000, 2005, 2010, and 2014 Cambodia Demographic and Health Survey (CDHS). The CDHS is a population-based household survey that is regularly carried out every 5 years. It is nationally representative. Which uses two-stage stratified cluster sampling to collect the samples from all provinces that are divided into sampling domains. They were further divided into sampling strata between urban and rural. In the first stage, cluster, or enumeration areas (EAs), that represents the entire country (urban and rural) are randomly selected from the sampling frame using probability proportional (PPS) to cluster size. In the second stage, a complete listing of households was selected from each cluster chosen using an equal probability systematic sampling, and then interviews with women between the aged 15-49 years who were born in the five years preceding the survey in the full list selected households. Details of CDHS design and data collection procedures have been described elsewhere [8-10]. Then, we limited our analysis to children born in the last five years prior to the surveys, alive and living with their mothers or caregivers during interview time.

### MEASUREMENTS

#### Outcome variable

Acute Respiratory Infection (ARI) is the outcome variable used in the study. CDHS was collected of ARI symptoms among children under the age of 5 years as the occurrence of cough accompanied by short, rapid breathing in the two weeks preceding the survey. Then, the outcome variable was then thought of as a binary variable, with a code of **1** indicating ARI and **0** otherwise.

#### Independent variables

Mother’s **Age** was categorized into 15-19 (reference group), 20-24, 25-29, 30-34, 35-39, and 40-49. Mother’s **Education** was coded into ordinal level variable with no education (reference group), primary, secondary, and higher. Mother’s **Employment** and Mother’s **Smoking** cigarettes were coded into a dichotomous variable with Not working vs Working; Not smoking vs Non-smoking. Child’s **Age** in months was coded as 0-5 (reference group), 6-11, 12-23, 24-35, 36-47, and 48-59. Children’s **Birth order** was coded into ordinal variable with 1^st^ child (reference group), 2-3 children, and at less 4 children. Child’s **Sex**, and **Weight’s at birth** in kilogram were coded into dichotomous variable with Less than 2.5kg vs 2.5kg and above. Also, **BCG vaccination status** was coded in to dichotomous variable with Incomplete vs Completed. **Received vitamin A last 6 months**, and **Breastfeeding children. Places of delivery** was categorized: Public facilities (reference group), Private facilities and at home). **Households wealth quintile** were calculated scores based on household assets (television, bicycle/car, size of agricultural land, quantity of livestock), and dwelling characteristics (sources of drinking water, sanitation facilities, and materials used for constructing houses) using principal component analysis (PCA), and the scores given into five categories of wealth quintile (poorest, poorer, medium, richer, and richest) each comprising 20% of the population [8-10]. **Improved drinking water** options included rainwater, piped into the home, piped into the yard or plot, public taps or standpipes, tubed wells or boreholes, and protected wells and springs. The classification of other sources as non-improved. **Toilet facilities** that have ventilated/improved latrines or other types of toilets are considered improved, whereas those that don’t have any toilets are considered unimproved. **Residence** areas (urban vs rural). **CDHS survey years** was categorized 2000 (reference group), 2005, 2010, and 2014.

### STATISTICAL ANALYSIS

All statistical analysis performed by STATA version 16 (Stata Corp 2019, College Station, TX) The complex survey design was declared to be taken into account using the STATA command “survey” package, and all estimations were carried out using the survey-specific command “svy” using the standard sampling weight (v005/1,000,000), clustering, and stratification variables that were provided by DHS. ARI symptoms trends over time for the CDHS in 2000, 2005, 2010, and 2014 are presented first. Then, bivariate chi-square tests were used to determine whether there were any statistically significant relationships between the independent variables of interest, such as the socio-demographics of the mother and her offspring, household traits, and ARI symptoms. The multiple logistic regression model (at p-value 0.15) included any covariates that reached a significant level [16-18]. However, regardless of their level of significance, background variables like the mother’s age group, the child’s age, the child’s residence, and survey years were automatically included. To examine the magnitude of the unadjusted associations between ARI symptoms and mother and child socio-demographics and household characteristics, we pooled data from 2000 to 2014 and used simple logistic regression. To forecast the independent variable, multiple logistic regression was used.

## ETHNICAL DECLARATIONS

The CDHS was approved by the Cambodia National Ethics Committee for Health Research and the Institutional Review Board (IRB) of ICF in Rockville, Maryland, USA. The CDHS data are publicly accessible and were made available to the researchers upon request to the DHS Program, ICF at https://www.dhsprogram.com

## RESULTS

### Trends of ARI symptoms among children age 0-59 months

**Table 1** presents the distribution of ARI symptoms among children under five across the survey year. A total of 30,040 children under 5 years old were analyzed, included 7,828, 7,621, 7,727, and 6,864 from 2000, 2005, 2010 and 2014 respectively. Overall, trends of children age 0-59 months having ARI symptoms in the previous two weeks were declined from 19.9% [95% CI: 18.4-21.5] in 2000 to 8.6% [95% CI: 7.7-9.6] in 2005 to 6.4% [95% CI: 5.6-7.4] in 2010 then, decreased to 5.5% [95% CI: 4.8-6.3%] in 2014, it was statistically significant at p value **< 0**.**001**. When, stratified by age, prevalence of children with ARI symptoms peaked in 2000 at around 30% between ages 5-20 months and then gradually declined to about 10% in 2005, 2010, and 2014 (**Fig 1**).

**Table 1.** Trends ARI symptoms of Cambodia children aged between 0-59 months (2000-2014 CDHS)

| Variable | 2000 (n=7,828) |  | 2005 (n=7,621) |  | 2010 (n=7,727) |  | 2014 (n=6,864) |  | P-value |
| --- | --- | --- | --- | --- | --- | --- | --- | --- | --- |
|  | % | 95%CI | % | 95%CI | % | 95%CI | % | 95%CI |  |
| ARI symptoms past two weeks |  |  |  |  |  |  |  |  |  |
| Yes | 19.9 | [18.4-21.5] | 8.6 | [7.7-9.6] | 6.4 | [5.6-7.4] | 5.5 | [4.8-6.3] | <0.001 |
| No | 80.1 | [78.5-81.6] | 91.4 | [90.4-92.3] | 93.6 | [92.6-94.4] | 94.5 | [93.7-95.2] |  |

### Association with ARI symptoms in children ages 0-59 months in bivariate analysis

Describes study population (**Table 2**). Mother’s aged 15-19 years old were 22.9%, 26.1%, 24.7%, and 26.3% from 2000, 2005, 2010 and 2014. While mothers have no education 37.4% in 2000, 28.8% in 2005, 21.6% in 2010, only 12.5% in 2014. Most of the mother had smoking the cigarette were 32.6%, 36.8%, 16.9% and 13.7%. Proportion of children were categorized by aged between 22-29% each age groups.

**Table 2.**
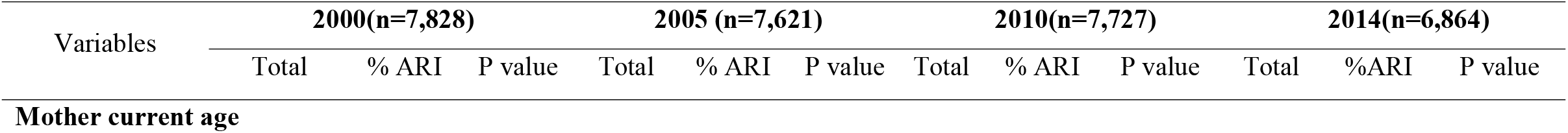
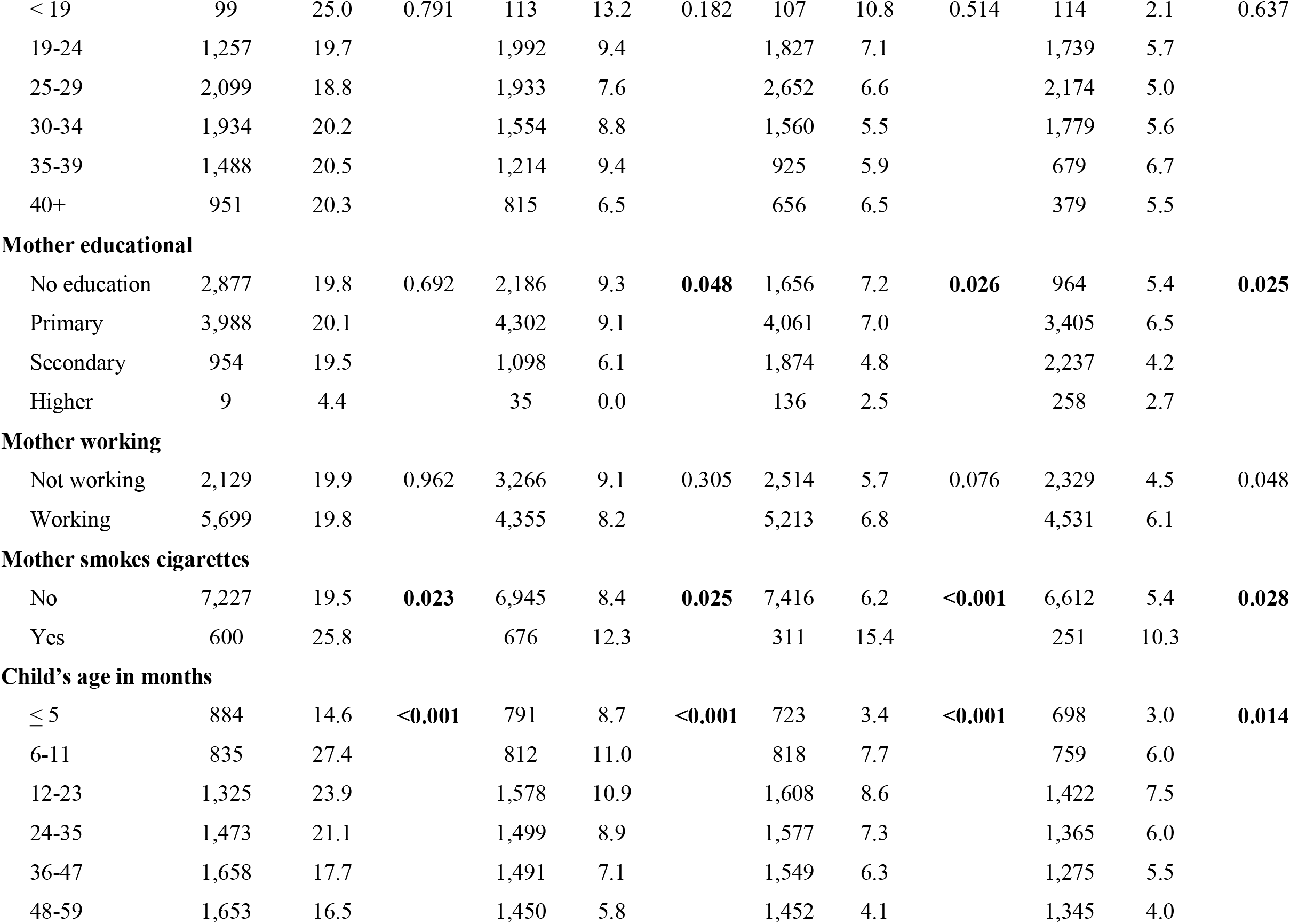
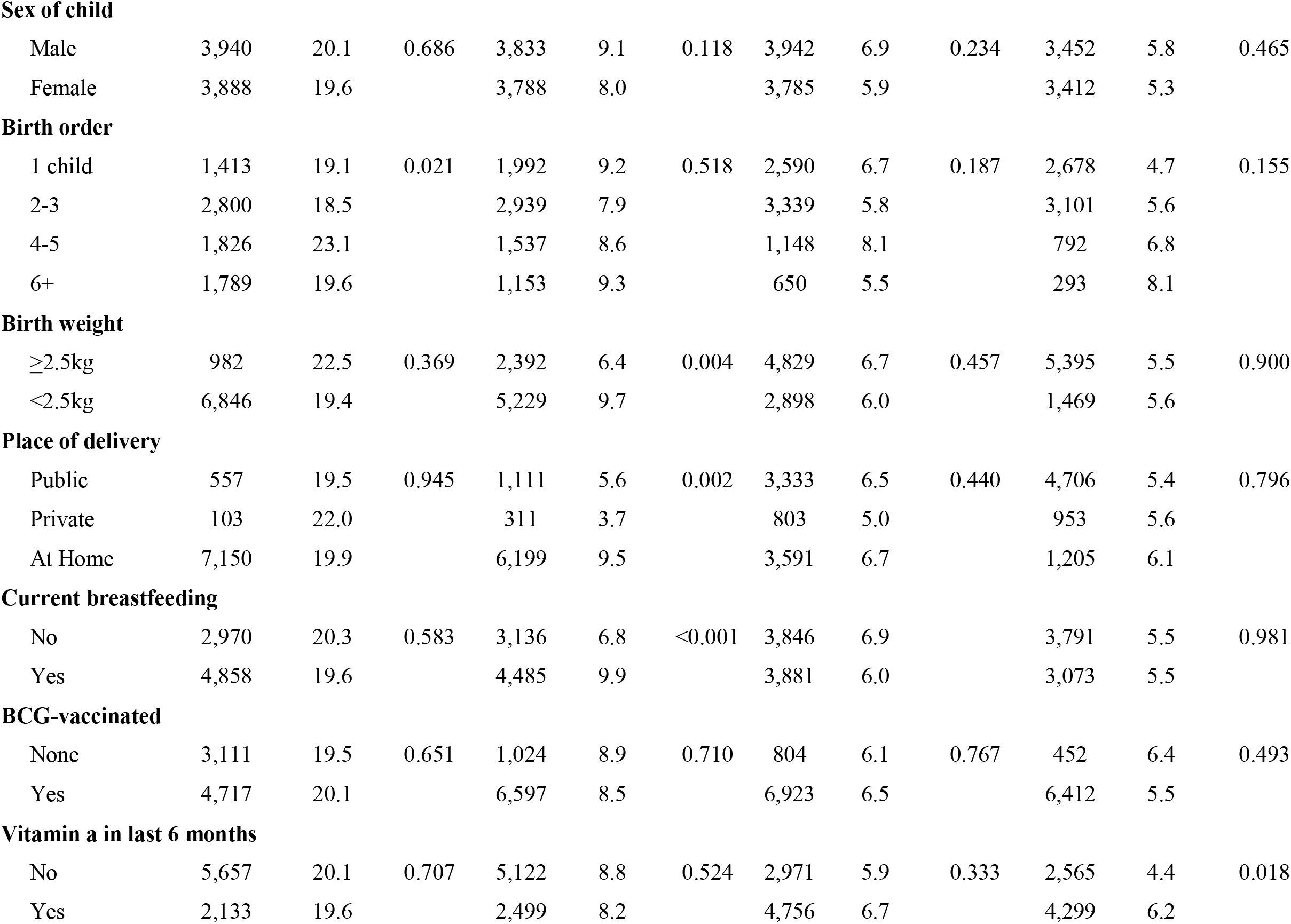
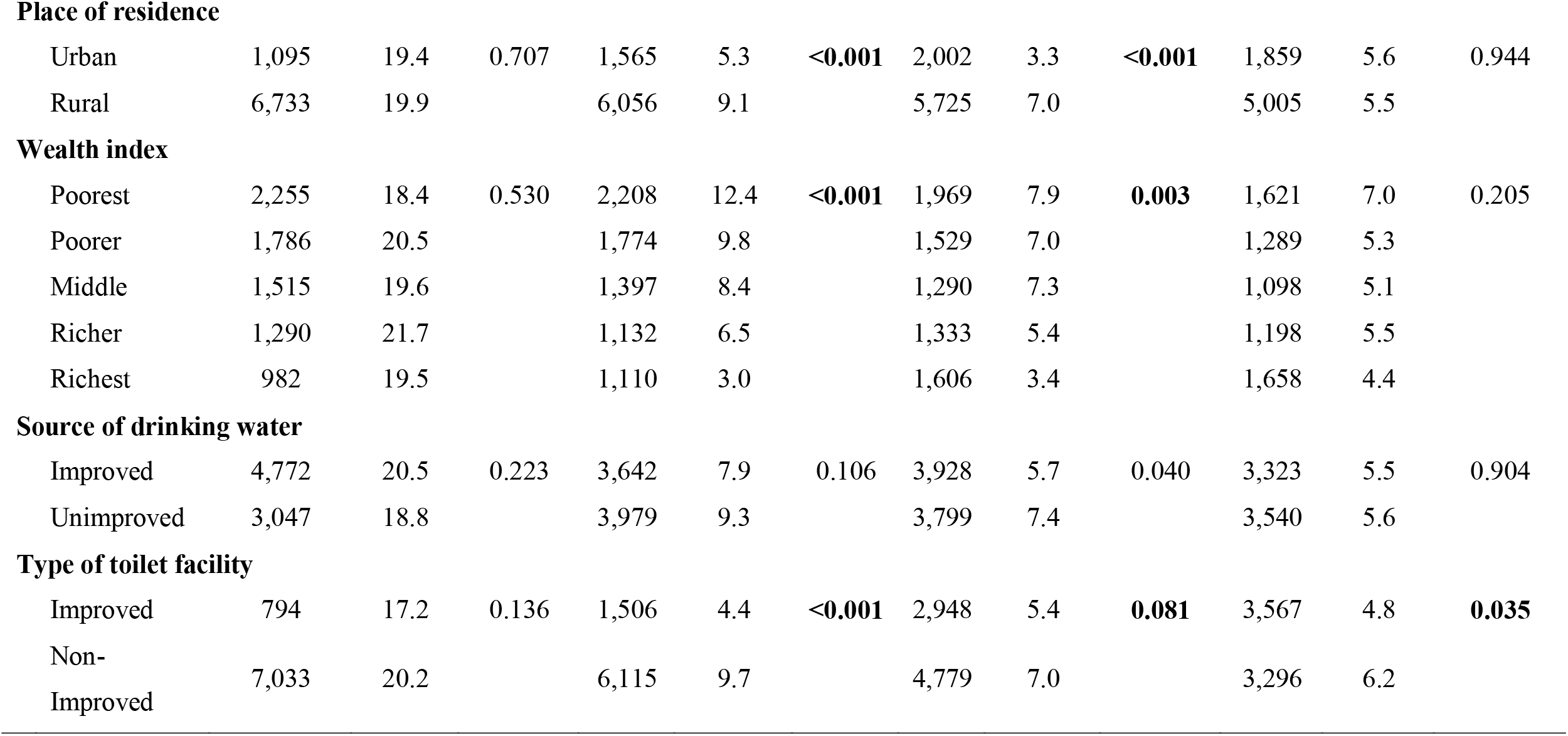
Proportion of children age 0-59 months who have ARI symptoms over survey years by characteristics in bivariate chi-square analysis, CDHS 2000,2005, 2010 and 2014

| Variables | 2000(n=7,828) |  |  | 2005 (n=7,621) |  |  | 2010(n=7,727) |  |  | 2014(n=6,864) |  |  |
| --- | --- | --- | --- | --- | --- | --- | --- | --- | --- | --- | --- | --- |
|  | Total | % ARI | P value | Total | % ARI | P value | Total | % ARI | P value | Total | %ARI | P value |
| **Mother current age** |  |  |  |  |  |  |  |  |  |  |  |  |
| < 19 | 99 | 25.0 | 0.791 | 113 | 13.2 | 0.182 | 107 | 10.8 | 0.514 | 114 | 2.1 | 0.637 |
| 19-24 | 1,257 | 19.7 |  | 1,992 | 9.4 |  | 1,827 | 7.1 |  | 1,739 | 5.7 |  |
| 25-29 | 2,099 | 18.8 |  | 1,933 | 7.6 |  | 2,652 | 6.6 |  | 2,174 | 5.0 |  |
| 30-34 | 1,934 | 20.2 |  | 1,554 | 8.8 |  | 1,560 | 5.5 |  | 1,779 | 5.6 |  |
| 35-39 | 1,488 | 20.5 |  | 1,214 | 9.4 |  | 925 | 5.9 |  | 679 | 6.7 |  |
| 40+ | 951 | 20.3 |  | 815 | 6.5 |  | 656 | 6.5 |  | 379 | 5.5 |  |
| **Mother educational** |  |  |  |  |  |  |  |  |  |  |  |  |
| No education | 2,877 | 19.8 | 0.692 | 2,186 | 9.3 | **0.048** | 1,656 | 7.2 | **0.026** | 964 | 5.4 | **0.025** |
| Primary | 3,988 | 20.1 |  | 4,302 | 9.1 |  | 4,061 | 7.0 |  | 3,405 | 6.5 |  |
| Secondary | 954 | 19.5 |  | 1,098 | 6.1 |  | 1,874 | 4.8 |  | 2,237 | 4.2 |  |
| Higher | 9 | 4.4 |  | 35 | 0.0 |  | 136 | 2.5 |  | 258 | 2.7 |  |
| **Mother working** |  |  |  |  |  |  |  |  |  |  |  |  |
| Not working | 2,129 | 19.9 | 0.962 | 3,266 | 9.1 | 0.305 | 2,514 | 5.7 | 0.076 | 2,329 | 4.5 | 0.048 |
| Working | 5,699 | 19.8 |  | 4,355 | 8.2 |  | 5,213 | 6.8 |  | 4,531 | 6.1 |  |
| **Mother smokes cigarettes** |  |  |  |  |  |  |  |  |  |  |  |  |
| No | 7,227 | 19.5 | **0.023** | 6,945 | 8.4 | **0.025** | 7,416 | 6.2 | **<0.001** | 6,612 | 5.4 | **0.028** |
| Yes | 600 | 25.8 |  | 676 | 12.3 |  | 311 | 15.4 |  | 251 | 10.3 |  |
| **Child's age in months** |  |  |  |  |  |  |  |  |  |  |  |  |
| ≤ 5 | 884 | 14.6 | **<0.001** | 791 | 8.7 | **<0.001** | 723 | 3.4 | **<0.001** | 698 | 3.0 | **0.014** |
| 6-11 | 835 | 27.4 |  | 812 | 11.0 |  | 818 | 7.7 |  | 759 | 6.0 |  |
| 12-23 | 1,325 | 23.9 |  | 1,578 | 10.9 |  | 1,608 | 8.6 |  | 1,422 | 7.5 |  |
| 24-35 | 1,473 | 21.1 |  | 1,499 | 8.9 |  | 1,577 | 7.3 |  | 1,365 | 6.0 |  |
| 36-47 | 1,658 | 17.7 |  | 1,491 | 7.1 |  | 1,549 | 6.3 |  | 1,275 | 5.5 |  |
| 48-59 | 1,653 | 16.5 |  | 1,450 | 5.8 |  | 1,452 | 4.1 |  | 1,345 | 4.0 |  |
| **Sex of child** |  |  |  |  |  |  |  |  |  |  |  |  |
| Male | 3,940 | 20.1 | 0.686 | 3,833 | 9.1 | 0.118 | 3,942 | 6.9 | 0.234 | 3,452 | 5.8 | 0.465 |
| Female | 3,888 | 19.6 |  | 3,788 | 8.0 |  | 3,785 | 5.9 |  | 3,412 | 5.3 |  |
| **Birth order** |  |  |  |  |  |  |  |  |  |  |  |  |
| 1 child | 1,413 | 19.1 | 0.021 | 1,992 | 9.2 | 0.518 | 2,590 | 6.7 | 0.187 | 2,678 | 4.7 | 0.155 |
| 2-3 | 2,800 | 18.5 |  | 2,939 | 7.9 |  | 3,339 | 5.8 |  | 3,101 | 5.6 |  |
| 4-5 | 1,826 | 23.1 |  | 1,537 | 8.6 |  | 1,148 | 8.1 |  | 792 | 6.8 |  |
| 6+ | 1,789 | 19.6 |  | 1,153 | 9.3 |  | 650 | 5.5 |  | 293 | 8.1 |  |
| **Birth weight** |  |  |  |  |  |  |  |  |  |  |  |  |
| ≥2.5kg | 982 | 22.5 | 0.369 | 2,392 | 6.4 | 0.004 | 4,829 | 6.7 | 0.457 | 5,395 | 5.5 | 0.900 |
| <2.5kg | 6,846 | 19.4 |  | 5,229 | 9.7 |  | 2,898 | 6.0 |  | 1,469 | 5.6 |  |
| **Place of delivery** |  |  |  |  |  |  |  |  |  |  |  |  |
| Public | 557 | 19.5 | 0.945 | 1,111 | 5.6 | 0.002 | 3,333 | 6.5 | 0.440 | 4,706 | 5.4 | 0.796 |
| Private | 103 | 22.0 |  | 311 | 3.7 |  | 803 | 5.0 |  | 953 | 5.6 |  |
| At Home | 7,150 | 19.9 |  | 6,199 | 9.5 |  | 3,591 | 6.7 |  | 1,205 | 6.1 |  |
| **Current breastfeeding** |  |  |  |  |  |  |  |  |  |  |  |  |
| No | 2,970 | 20.3 | 0.583 | 3,136 | 6.8 | <0.001 | 3,846 | 6.9 |  | 3,791 | 5.5 | 0.981 |
| Yes | 4,858 | 19.6 |  | 4,485 | 9.9 |  | 3,881 | 6.0 |  | 3,073 | 5.5 |  |
| **BCG-vaccinated** |  |  |  |  |  |  |  |  |  |  |  |  |
| None | 3,111 | 19.5 | 0.651 | 1,024 | 8.9 | 0.710 | 804 | 6.1 | 0.767 | 452 | 6.4 | 0.493 |
| Yes | 4,717 | 20.1 |  | 6,597 | 8.5 |  | 6,923 | 6.5 |  | 6,412 | 5.5 |  |
| **Vitamin a in last 6 months** |  |  |  |  |  |  |  |  |  |  |  |  |
| No | 5,657 | 20.1 | 0.707 | 5,122 | 8.8 | 0.524 | 2,971 | 5.9 | 0.333 | 2,565 | 4.4 | 0.018 |
| Yes | 2,133 | 19.6 |  | 2,499 | 8.2 |  | 4,756 | 6.7 |  | 4,299 | 6.2 |  |
| **Place of residence** |  |  |  |  |  |  |  |  |  |  |  |  |
| Urban | 1,095 | 19.4 | 0.707 | 1,565 | 5.3 | **<0.001** | 2,002 | 3.3 | **<0.001** | 1,859 | 5.6 | 0.944 |
| Rural | 6,733 | 19.9 |  | 6,056 | 9.1 |  | 5,725 | 7.0 |  | 5,005 | 5.5 |  |
| **Wealth index** |  |  |  |  |  |  |  |  |  |  |  |  |
| Poorest | 2,255 | 18.4 | 0.530 | 2,208 | 12.4 | **<0.001** | 1,969 | 7.9 | **0.003** | 1,621 | 7.0 | 0.205 |
| Poorer | 1,786 | 20.5 |  | 1,774 | 9.8 |  | 1,529 | 7.0 |  | 1,289 | 5.3 |  |
| Middle | 1,515 | 19.6 |  | 1,397 | 8.4 |  | 1,290 | 7.3 |  | 1,098 | 5.1 |  |
| Richer | 1,290 | 21.7 |  | 1,132 | 6.5 |  | 1,333 | 5.4 |  | 1,198 | 5.5 |  |
| Richest | 982 | 19.5 |  | 1,110 | 3.0 |  | 1,606 | 3.4 |  | 1,658 | 4.4 |  |
| **Source of drinking water** |  |  |  |  |  |  |  |  |  |  |  |  |
| Improved | 4,772 | 20.5 | 0.223 | 3,642 | 7.9 | 0.106 | 3,928 | 5.7 | 0.040 | 3,323 | 5.5 | 0.904 |
| Unimproved | 3,047 | 18.8 |  | 3,979 | 9.3 |  | 3,799 | 7.4 |  | 3,540 | 5.6 |  |
| **Type of toilet facility** |  |  |  |  |  |  |  |  |  |  |  |  |
| Improved | 794 | 17.2 | 0.136 | 1,506 | 4.4 | **<0.001** | 2,948 | 5.4 | **0.081** | 3,567 | 4.8 | **0.035** |
| Non-Improved | 7,033 | 20.2 |  | 6,115 | 9.7 |  | 4,779 | 7.0 |  | 3,296 | 6.2 |  |

**Figure 1.**
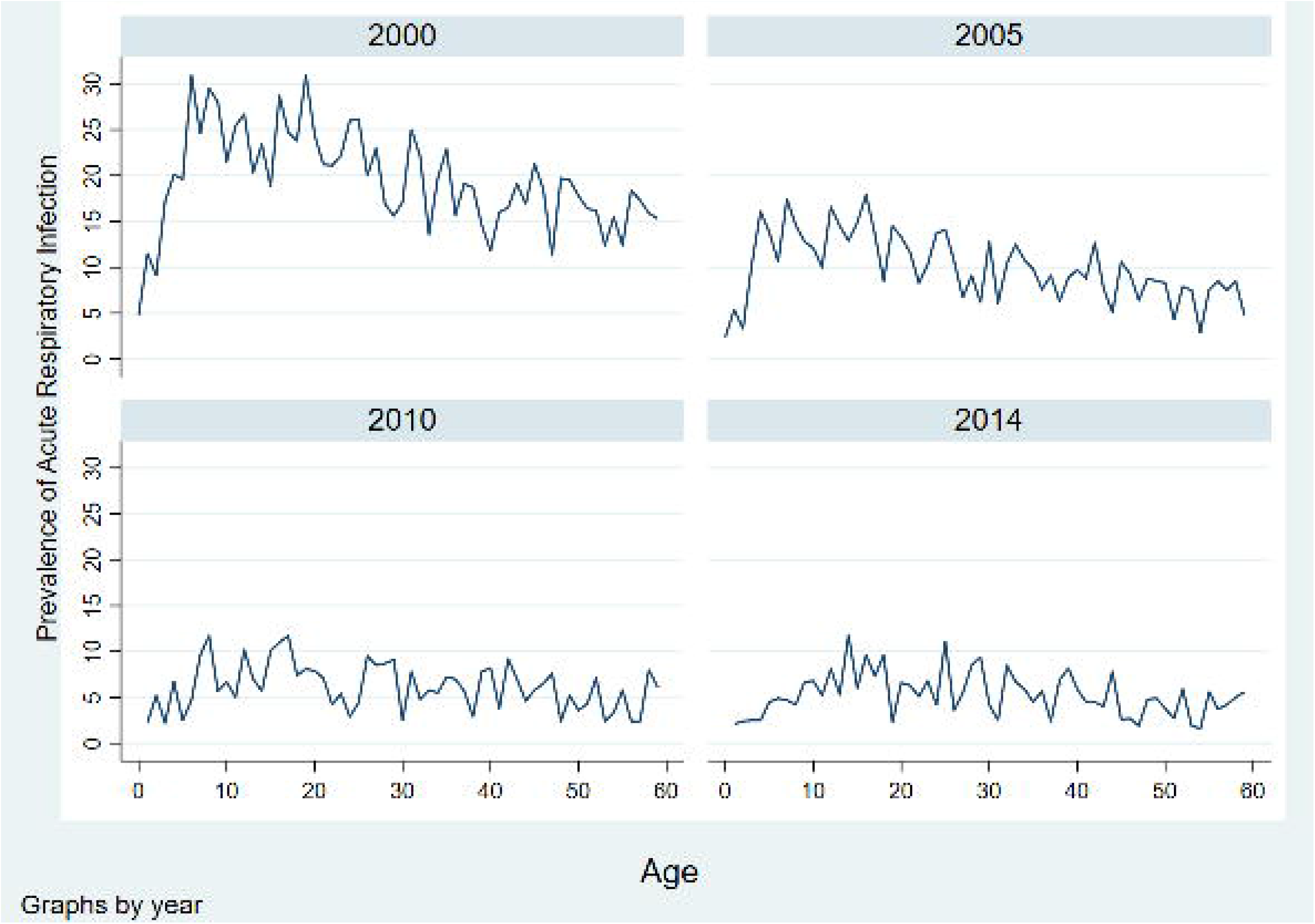
Trends of children aged 0-59 months with ARI symptoms by survey years, CDHS 2000,2005, 2010 and 2014.

In bivariate analysis of the data for children reveals that, with the exception of 2000, the proportion of children who had ARI symptoms in the previous two weeks was related to mothers’ educational attainment. In comparison to children from mothers with limited education, children from mothers with higher education had the lowest percentage of ARI symptoms (4.4%, 2.5%, and 2.7%, respectively) across all surveys. Similar to this, mothers who smoked cigarettes were more likely to have children with ARI symptoms than mothers who did not smoke (26% in 2000 at p value 0.023, 12.3% in 2005 at p value 0.025, 15.4% in 2010 at p value 0.001, and 17.5% at p value 0.001). ARI symptoms were related to children’s ages (6-11 months, 27.4% in 2000, 11.0% in 2005, 8% in 2010, and 6% in 2014; 12-23 months, 24%, 11%, 8.6%, and 7.5%, respectively; p values 0.001 in 2000-2010 and 0.014 in 2014). In children born into the richest household quantile, the prevalence of ARI symptoms was the lowest, ranging from 3% to 4.4% from 2005 to 2014, but 19.5% in 2000; at p values 0.001 in 2005 and 2010 (**Table 2**).

### Determinants of ARI symptoms in Multiple logistic analysis

In the final multiple logistic model (**Table 3**), factors independently increased likelihood that children would develop ARI symptoms included, children’s ages, mother’s smoking cigarette, household unimproved latrines. While mothers with higher education, breastfeeding children, children born into the wealthiest families, and survey years all reduced the likelihood of an ARI symptoms.

**Table 3.** Factors independently associated with ARI symptoms among children aged 0-59 months in multiple logistic regression analysis, CDHS 2000,2005, 2010 and 2014

| Variables | 2000-2014 (N=30,040) |  | (2000-2014) N=29,988 |  |
| --- | --- | --- | --- | --- |
|  | OR | 95% CI | AOR | 95% CI |
| **Mother current age** |  |  |  |  |
| < 19 | 1.0 | ref | 1.0 | ref |
| 19-24 | 0.74 | (0.51 - 1.07) | 0.79 | (0.54 - 1.16) |
| 25-29 | 0.70* | (0.48 - 1.01) | 0.73 | (0.49 - 1.07) |
| 30-34 | 0.79 | (0.55 - 1.14) | 0.74 | (0.50 - 1.11) |
| 35-39 | 0.93 | (0.64 - 1.36) | 0.77 | (0.50 - 1.19) |
| 40+ | 0.86 | (0.59 - 1.26) | 0.71 | (0.46 - 1.10) |
| **Mother working** |  |  |  |  |
| Not working | 1.0 | ref | 1.0 | ref |
| Working | 1.14** | (1.01 - 1.28) | 1.03 | (0.91 - 1.16) |
| **Mother educational** |  |  |  |  |
| No education | 1.0 | ref | 1.0 | ref |
| Primary | 0.86** | (0.76 - 0.98) | 1.09 | (0.95 - 1.23) |
| Secondary | 0.57*** | (0.48 - 0.68) | 0.93 | (0.77 - 1.13) |
| Higher | 0.18*** | (0.09 - 0.37) | **0.45\*\*** | **(0.21 - 0.94)** |
| **Mother smokes cigarettes** |  |  |  |  |
| No | 1.0 | ref | 1.0 | ref |
| Yes | 1.96*** | (1.58 - 2.43) | **1.61\*\*\*** | **(1.27 - 2.05)** |
| **Child age in months** |  |  |  |  |
| ≤ 5 | 1.0 | ref | 1.0 | ref |
| 6-11 | 1.81*** | (1.46 - 2.25) | **1.91\*\*\*** | **(1.53 - 2.38)** |
| 12-23 | 1.68*** | (1.39 - 2.04) | **1.79\*\*\*** | **(1.46 - 2.20)** |
| 24-35 | 1.45*** | (1.18 - 1.77) | **1.41\*\*\*** | **(1.13 - 1.76)** |
| 36-47 | 1.24** | (1.02 - 1.52) | 1.15 | (0.92 - 1.44) |
| 48-59 | 1.02 | (0.84 - 1.25) | 0.94 | (0.76 - 1.18) |
| **Sex of child** |  |  |  |  |
| Male | 1.0 | ref | 1.0 | ref |
| Female | 0.92* | (0.84 - 1.01) | 0.92* | (0.83 - 1.01) |
| **Birth order** |  |  |  |  |
| 1 child | 1.0 | ref | 1.0 | ref |
| 2-3 | 1.05 | (0.93 - 1.19) | 0.94 | (0.83 - 1.07) |
| 4-5 | 1.58*** | (1.37 - 1.81) | 1.14 | (0.95 - 1.38) |
| 6+ | 1.62*** | (1.39 - 1.90) | 0.97 | (0.77 - 1.22) |
| **Birth weight** |  |  |  |  |
| ≥ 2.5 Kg | 1.0 | ref | 1.0 | ref |
| < 2.5 Kg | 1.82*** | (1.61 - 2.05) | 0.87* | (0.74 - 1.01) |
| **Place of delivered** |  |  |  |  |
| Public | 1.0 | ref | 1.0 | ref |
| Private | 0.90 | (0.69 - 1.18) | 1.05 | (0.79 - 1.39) |
| At Home | 2.04*** | (1.78 - 2.33) | 1.11 | (0.93 - 1.33) |
| **Current breastfeeding** |  |  |  |  |
| No | 1.0 | ref | 1.0 | ref |
| Yes | 1.17*** | (1.06 - 1.30) | **0.87\*\*** | **(0.77 - 0.98)** |
| **BCG-vaccinated** |  |  |  |  |
| None | 1.0 | ref | 1.0 | ref |
| Yes | 0.61*** | (0.54 - 0.70) | 1.02 | (0.89 - 1.18) |
| **Vitamin a in last 6 months** |  |  |  |  |
| No | 1.0 | ref | 1.0 | ref |
| Yes | 0.75*** | (0.67 - 0.83) | 1.04 | (0.93 - 1.17) |
| **Place of residence** |  |  |  |  |
| Urban | 1.0 | ref | 1.0 | ref |
| Rural | 1.33*** | (1.10 - 1.59) | 1.04 | (0.86 - 1.24) |
| **Wealth index** |  |  |  |  |
| Poorest | 1.0 | ref | 1.0 | ref |
| Poorer | 0.94 | (0.82 - 1.07) | 0.93 | (0.82 - 1.07) |
| Middle | 0.88 | (0.76 - 1.03) | 0.89 | (0.76 - 1.04) |
| Richer | 0.86* | (0.72 - 1.01) | 0.90 | (0.75 - 1.07) |
| Richest | 0.57*** | (0.46 - 0.69) | **0.73\*\*** | **(0.56 - 0.95)** |
| **Type of toilet facility** |  |  |  |  |
| Improved | 1.0 | ref | 1.0 | ref |
| Non-Improved | 2.09*** | (1.80 - 2.42) | **1.20\*** | **(0.99 - 1.46)** |
| **Source of drinking water** |  |  |  |  |
| Improved | 1.0 | ref | - | - |
| Unimproved | 0.94 | (0.83 - 1.06) | - | - |
| **Survey years** |  |  |  |  |
| 2000 | 1.0 | ref | 1.0 | ref |
| 2005 | 0.38*** | (0.32 - 0.44) | **0.36\*\*\*** | **(0.31 - 0.42)** |
| 2010 | 0.28*** | (0.23 - 0.33) | **0.27\*\*\*** | **(0.22 - 0.33)** |
| 2014 | 0.24*** | (0.20 - 0.28) | **0.24\*\*\*** | **(0.19 - 0.30)** |

\*\*\* $p < 0.01$ , \*\* $p < 0.05$ , \* $p < 0.1$

Children were more likely to have ARI symptoms if they were aged 6-11 months [AOR = 1.91; 95% CI: 1.53-2.38], 12-23 months [AOR = 1.79; 95% CI: 1.46-2.20], 24-35 months [AOR = 1.41; 95% CI: 1.13-1.76] compared to children aged 0-5 months. Maternal smoking status had a significant effect on children’s ARI symptoms; the effects were significantly higher odds [AOR = 1.61; 95% CI: 1.27-2.05] among those born to mothers who smoked. In addition, children of households which unimproved toilet facility were associated with ARI symptoms [AOR = 1.20; 95% CI:0.99-1.46] compared to improved latrine. On the contrary, the following factors were independently associated with decreased odds of having ARI symptoms: Mother with higher education [AOR = 0.45; 95% CI: 0.21-0.94] compared to mothers with limited education. Children breastfeeding [AOR = 0.87; 95% CI: 0.77-0.98] compared to non-breastfeeding. Children born into richest wealth quantile [AOR = 0.73; 95% CI: 0.56-0.95] compared to poorest wealth quantile. Lastly, Survey years in 2005 [AOR = 0.36; 95% CI: 0.31 - 0.42], 2010 [AOR = 0.27; 95% CI: 0.22 - 0.33], 2014 [AOR = 0.24; 95% CI: 0.19 - 0.30] compared in 2000 (**Table 3**).

## DISCUSSION

Prevalence of ARI among children U5 in Cambodia highly decreased from 19.5% in 2000 to 5.2% in 2014. These findings agree with global efforts to reduce under five mortality and efforts to end child deaths due to ARI and diarrhea [7, 15, 19]. The reduction that was noted in the prevalence of ARI can be attributed to the efforts by the Cambodian National Immunization Program’s efforts to replace the DPT vaccine with a tetravalent vaccine that includes DPT and the Hib vaccine and a pentavalent vaccine that includes the DPT, Hib, and hepatitis B vaccine are responsible for the decrease in the prevalence of ARI that was observed (HepB). The HepB vaccine is also given as part of the program in 2006 [10], either at birth or during the first clinical encounter. Additionally, from 40% in 2000 to 96% in 2014, more children received the BCG vaccine [10].

Children who are exposed to their mothers’ smoking have odds 1.6 times greater chance of developing ARI symptoms. This study’s findings regarding the link between a mother’s smoking and ARI symptoms were consistent with those of studies done in other sub-Saharan African nations [6, 20-22]. Children who are exposed to parental smoking are more likely to have pneumonia and other respiratory infection diseases, according to WHO reports [23]. Particularly in areas where smoking and the use of firewood are common, parents and the community need to be educated about the risks that smoking has for children [24, 25]. ARI symptoms are more common in young children (0-35 months) than in older children (36-59 months). These results are in line with other studied [12, 24, 26] that indicated an increased likelihood of ARI. The increased risk for ARI in this age group is probably due to the children’s low immunity, which tends to increase as results of exposures to vaccination and generally development of infections resistance. In particularly in countries in Southeast Asia and sub-Saharan African, where health facilities and maternal healthcare education need to be improved, the factors were low rates of immunization in young children, low maternal literacy, and young mothers engaged in farming activities that prevent the care of young children [26, 27]. Additionally, compared to households with improved toilet facilities, children from households with unimproved toilet facilities are more likely to experience ARI symptoms. Improved sanitation facilities reduced the risk of fever by 13% and the risk of cough by 10%, according to a supporting multicounty WASH-intervention study; however, the reduction in childhood mortality was very low [28]. This study suggests that child infection diseases are all related to the lack of all three WASH facilities. Similar studies done in Nigeria and Myanmar found that households lacking all three types of WASH facilities had higher odds of having cough, fever, and diarrhea [29, 30]. Inversely, regarding mother behavior factors, such as breastfeeding, which conflicts with other findings [31, 32]. According to the current study, non-breastfed children were found to be more likely than breastfed children to experience the symptoms of ARI disease. In general, breastfeeding is more crucial for a child’s nutrition and the health of their immune system. Similarly, children whose mothers had a secondary or higher education were less likely than children whose mothers had no education to experience ARI symptoms. These results are in line with research done in Kenya, Ethiopia, and Rwanda [11, 33, 34]. This finding might be explained by the fact that these mothers might have access to books that give them advice on how to safeguard their children. Additionally, this research showed that children in higher wealth quintile households had a lower risk of developing ARI symptoms. Consistent with studied conducted in Bangladesh [35]. Higher family income has been linked to better living conditions, better nutritional status, and access to healthcare services, all of which have a positive impact on children’s health outcomes. Financial stress on parents has a variety of effects on children’s health and susceptibility to disease. For example, undernutrition, impaired cognitive development, and a weakened immune system in children are all strongly associated with financial stress, which increases the risk of infectious diseases. Compared to children from poor families, those from financially stable families are more likely to enjoy safe and secure housing with greater access to health-promoting conditions [36]. Children who were gathered between 2005 2010, and 2014 had a lower risk of developing ARI symptoms. Confirmed by studied conducted in East and South-East Asian nations [37]. This decrease in the burden of ARI in these developing regions is the result of both a decline in incidence brought on by socioeconomic development and higher living standards as well as a rise in access and quality of care [19]. While proportion of children age 12-23 months who have been fully vaccinated against all basic antigens lower at 40% to 67% between 2000-2005 and peaked at 79% in 2010, has declined to 73% in 2014, and slightly increased to 76% in 2021-22 [38]. This study also demonstrated that, compared to 2000, there have been notable advancements in reducing U5 mortality due to infection diseases, which has been reduced, and achieved some remarkable child health outcomes reported to Millennium Development Goals (MDGs), such as a significant decline in child mortality rates in Cambodia by 2015 [39].

The study has the number of limitations. First, the CDHS was a cross-sectional study, the temporal associations, or causality between associated independent variables with ARI symptoms could not unable to assess at the same times. In addition, our study used secondary data from four different surveys such as the effect of some variables noted in literature were not assessed for example, the effect of children malnutritional, type of roofing material, household’s types of cooking, season effect, mode of delivered and number of antenatal care visits during pregnant due to there has much missing information were excluded variables from analysis. Secondly, the outcome variable ARI was based on self-report from the mothers, possible recall bias. From the newest CDHS data are publicly accessible and were made available to the researchers upon request to the DHS Program, ICF at https://www.dhsprogram.com four surveys round years were used for analysis. However, these data were gathered in 2000, 2005, 2010, and 2014 and might no longer reflect the current situation of children in Cambodia. Further evaluation of the prevalence of ARI symptoms and its determinants from CDHS 2020/21 could be done when the data becomes publicly available. Thirdly, the study was able to chart the trends and identify the factors associated with ARI among under five children and it is hoped that these findings will help the Cambodia Ministry of Health’s Child Health Unit as well as health promotion and social determinants to plan interventions which will contribute to the reduction of ARI symptoms. Despite the limitations, the study has strengths. The use of nationally representative data and a large sample size is a major strength of the study.

In conclusion, Cambodia have made highly decreased prevalence of ARI among children aged 0-59 months from 19.5% in 2000 to 5.2% in 2014. It agreed with global and Cambodia efforts to reduce under five deaths. This paper revealed that the mother’s smoking, child’s age, and households which unimproved toilet facilities were found to be potential risk factors for ARI symptoms in children under the age of 5 years. However, children living into mother education with richest wealth status, breastfed children had a significant reduce odds of the development of ARI disease symptoms. Based on the findings, policymakers and stakeholders in the health sector should launch targeted initiatives to address the issues with inadequate child healthcare, unfavorable environmental conditions, and childcare facilities. Government and child family programs must promote maternal education, particularly infant breastfeeding. The government ought to support maternal education and infant breastfeeding in the interest of early childhood care.

## Data Availability

The CDHS data used is openly available to the DHS Program website after having access approval at https://www.dhsprogram.com/data/

## ACKNOWLEDGMENTS

The authors would like to thank DHS-ICF, who approved the data used for this paper.

## AUTHORS’ CONTRIBUTIONS

**Um S**. contributed to conceptualization, methods, data analysis, and writing original draft, writing review & editing; **Pin P**. contributed to conceptualization, and writing original introduction draft. **Vang D**. contributed to writing review & editing manuscript; **Darapheak C**. contributed to writing review & editing. All authors read and approved the final manuscript.

## ABBREVIATIONS

ARI: Acute Respiratory Infections
WHO: World Health Organization
CDHS: Cambodia Demographic Health Survey
U5: Children Under Five
AOR: Adjusted odds ratio
PPS: Probability proportional to size
EA: Enumeration areas
BCG: Bacillus Calmette-Guérin
MDGs: Millennium Development Goals

